# Modulatory effect of Human immunodeficiency virus on serum miR-21, miR-125b and p53 gene: Any prognostic implication?

**DOI:** 10.1101/2022.12.28.22284013

**Authors:** Jude Ogechukwu Okoye, Anthony Ajuluchukwu Ngokere, Charles Chinedum Onyenekwe, Olaposi Idowu Omotuyi, Samuel Ifedioranma Ogenyi, Chioma Maureen Obi, Samuel Ayobami Fasogbon

**Affiliations:** Department of Medical Laboratory Science, Nnamdi Azikiwe University, Nnewi, Nigeria; Department of Medical Laboratory Science, Babcock University, Ilishan-Remo, Nigeria; Centre for Biocomputing and Drug Development, Adekunle Ajasin University, Department of Biochemistry and Biotechnology, Adekunle Ajasin University, Akungba, Ondo State; Medical Laboratory Science Council of Nigeria, Abuja, Nigeria

## Abstract

**Introduction:** Identifying individuals at a high risk of developing cervical cancer remains a major challenge, especially among individuals living with HIV. This study evaluated the levels of normally downregulated oncomirs (miR-21, miR-146a, miR-155, miR-182, and miR-200c) and normally upregulated tumor suppressors (miR-let-7b, miR-125b, miR-143, miR-145, and p53 expression) associated with cervical cancer in the serum of women living with and without HIV.

**Methods:** This case-control study was carried out between the months of May (2017) and April (2019). It included 173 women without abnormal Pap smears and negative for Human papillomavirus and Epstein-Barr virus; confirmed HIV seropositive women (HIV+; n= 103) and HIV seronegative women (HIV-; n= 70). Relative expressions of miRNAs, and the p53 gene in serum were determined using reverse transcriptase PCR and gel electrophoresis. T-test and Pearson’s correlation analyses were carried out on the generated data. Statistical significance was set at *p*≤ 0.05 and 0.01.

**Results:** A significantly higher level of miR-21 and lower levels of miR-125b and p53 gene were observed among HIV+ women compared with their HIV-counterparts at p= 0.028, 0.050, and 0.049, respectively. Among HIV+ women, significant direct correlations were observed between miR-21 and other oncomirs, including miR-145 (p< 0.05) while a significant inverse correlation was observed between miR-21 and miR-let-7b level (p= 0.013).

**Conclusion:** This study suggests that a high circulating level of miR-21 and a low circulating level of miR-125b could be used as biomarkers for identifying individuals at risk of developing cervical cancer, especially among HAART-naïve women who are living with HIV.

## Introduction

Globally, cervical cancer accounts for about 10% of all malignancies observed among women with a mortality rate of 49% among affected women.^1^ Despite the fact that the prevalence of Human immunodeficiency virus (HIV) reduced from 5.0% to 3.2% from 2003 to 2014, the negative impact of HIV on the trend of cervical cancer in Nigeria is still being felt.^2^ From 2005 to 2016, the incidence of the disease in Nigeria increased from 25 per to 28.3 per 10,000.^3,4^ As of 2018, reports show that the prevalence of cervical cancer attributable to HIV was higher in Africa than in other continents. Evidence shows that 25% of women living with HIV develop premalignant lesions and 12-30% of such women develop invasive cervical cancer despite receiving antiviral therapy.^5^ The virus has been known to promote the development of cervical cancer through CD4 T cell apoptosis or reduced clearance of onco-viruses.^5^ This suggests that HIV is a major player in epithelial carcinogenesis. In the face of immune suppression, viral infection elicits overwhelming dysregulation of the host’s gene and malignant epithelial transformation, mostly by manipulating microRNA–mRNA networks through microRNA stability and translation alteration. MicroRNA (miR) is a group of short non-coding RNAs that impedes gene expression through translational blocking or mRNA degradation.^6^ In the event of DNA damage, the p53 gene regulates cell proliferation, apoptosis, and senescence. Its mutation or degradation has been associated with cervical cancer.^7^ Identifying reliable biomarkers is vital for the effective monitoring of high-risk groups which would ultimately reduce late-stage presentation and cervical cancer-related death. This study evaluated the levels of normally downregulated oncomirs (miR-21, miR-146a, miR-155, miR-182, and miR-200c) and normally upregulated tumor suppressors (miR-let-7b, miR-125b, miR-143, miR-145, and p53 expression) associated with cervical cancer in the serum of women living with and without HIV in a bid to identify early indicators of genetic instability.^8,9^

## Materials and methods

### Ethical consideration and informed Consent

For this study, ethical approvals were obtained from the State Hospital Abeokuta Ethics Committee (SHA/RES/VOL.2/177) and Babcock University Health Research Committee (BUHREC549/18). As recommended by the ethics committee, written and signed informed consent was obtained from each participant before samples were collected. This case-control study was carried out between the months of May (2017) and April (2019). It included HIV uninfected women (HIV-; n= 70) and women living with HIV (HIV+; n= 103) living in Abeokuta.

### Sample collection, handling, and assays

Participants were screened for cervical cancer using a conventional Pap smear. Blood samples were collected at the Family Planning and HIV clinics at State Hospital Ijaiye, Ogun State. The separated sera were discharged into plain tubes, and stored at −20°C in the Department of Physiology, Babcock University, until they were analyzed. Five (5) ml of peripheral blood was also collected into an ethylenediaminetetraacetic acid vacutainer tube for CD4+ T-cell counts and analyzed within 2 hours of collection using the CD4 easy count kit and Cyflow Counter in Department of Chemical Pathology, Babcock University Teaching Hospital, Ogun State. Antibodies against HIV1/2 were tested according to the manufacturer’s instructions using ELISA kits (from Qingdao Hightop Biotech Co. Ltd, China) with positive and negative cut-off values for HIV-1/2 antibodies were 1.077 and 1.076, respectively. Individuals with any history of Human papillomavirus and Epstein-Barr virus infection, cancer, especially cervical, breast, and oral cancers were excluded.^9,10^

### RNA isolation from serum

The optimized phenol-chloroform method was used for RNA extraction in the Centre for Biocomputing and Drug Development, Adekunle Ajasin University.^10^ Fifty (50) μl of serum was added to Eppendorf tubes containing 50 μl trizol reagent and vortexed at 2500rpm for 15mins using a vortex mixer. The supernatants containing the RNA were aspirated into new labeled tubes. Iso-amyl alcohol (100 μL) was added to the supernatant with subsequent centrifugation for 30 min at 1500 revolutions per minute. For the cervical cells, 50 μl of Liquid-based Prep infranatant from each woman was homogenized in a different Eppendorf tube containing 50μl trizol reagent by pipetting up and down and later vortexed as well. Chloroform (100 μL) was added to the mixture which was subsequently vortexed and centrifuged for 30 min at 1500 revolutions per minute. The RNA was recovered in pellet form following decantation of the supernatant containing initially containing serum and cervical cells. The RNA pellet was washed thrice with 70% ethanol. Fifty μL of 70% ethanol was added to the tube and the tube was centrifuged for 5 min at 1500 revolutions per minute. After which, the supernatant was decanted. After washing all tubes were allowed to air dry. Fifty microlitres (μL) of nuclease-free water was added to the total RNA to form the RNA solution. The total RNA concentration (48 μL of deionized distilled water + 2 μL of RNA solution) was quantified using a spectrophotometer at 260 nanometres. The RNA limit of importance was set at 0.05–1.00. Acceptable RNA quality was set at 1.8-2.2 (based on OD260/OD280 calculation).^9,10^

### Complementary DNA synthesis

The cDNA synthesis was carried out by adding a miRNA-universal stem-loop primer cocktail (containing nuclease-free water, the reverse transcriptase buffer, the reverse transcriptase, miRNA-universal stem-loop primer specific oligos, and oligo deoxyribonucleotide triphosphate) to 20 μL of total RNA of each sample. The samples were then incubated at room temperature overnight. The concentration of complementary DNA was determined spectrophotometrically at 260 nanometres and homogeneity was carried out on every sample. To establish homogeneity of cDNA (HcDNA) concentration across all samples, the formula HcDNA= VB – VA was applied; where VB (dilution volume) = CA x VA/CB; CA= absorbance reading of RNA x 40 × 25, VA= needed volume of cDNA (25) and CB= least absorbance reading. All samples were diluted to the same concentration.^9,10^

### Reverse transcriptase polymerase chain reaction

The primers for miRNA quantification included: miR-Let-7b forward (5’-GTTTCGGGGTGAGG TAGTA-3’), miR-16 forward (5’-GTTGTCAGCAGTGCCTTAG-3’), miR-21 forward (5’-GG TGTCGGGTAGCTTATCA-3’), miR-125b forward (5’-GTTTTGCGCTCCTCTCAGT-3’), miR-143 forward (5’-TTTTTGCGCAGCGCCCTG-3’), miR-145 forward (5’-GTTTCACCTTGTC CTCACG-3’), miR-155 forward (5’-GTTTCTGTTAATGCTAATCGTGATA-3’), miR-182 forward (5’-GTTTTAGAACTCACACGTGTGA-3’), miR-200c forward (5’-GTTTCCCTCGT CTTACCCA-3’), universal reverse primer (5’-GTGCAGGGTCCGAGGT-3’), wild type p53 forward (5’-GCTCAAGACTGGCGCTAAAA-3’), wild type p53 reverse (5’-GTGACTCA GAGAGGACTCAT-3’); β-actin forward (5’-ACACTTTCTACAATGAGCTGCG-3’) β-actin reverse (5’-ACCAGAGGCATACAGGACAA C-3’) and miRNA-USLP (Universal stem-loop primer; 5’-AGTGCAGGGTCCGAG GTATTCGCACCAGAGCCAACATGTCACG-3’). The amplification was performed using optimization. Template (cDNA) 2μl, nuclease-free water 3μl, forward primers 0.5μl and reverse primers 0.5μl (Inqaba Biotechnical industries Ltd, South Africa) and master mix 4μl (Biolabs, South Africa). All reagents were added to each sample for a complete enzymatic reaction, and PCR was carried out. Amplification conditions were: 94°C pre-denaturation for 5mins, 94°C for 30 sec, annealing 55°C for 30sec and Extension 72°C for 30sec and then 5min at 72°C by 45 cycles).^9,10^

### Gel Electrophoresis

Products from PCR were electrophoresed in 0.5% of agarose gel using 0.5x TBE buffer of pH 8.3 (2.6g of Tris base, 5.0g of Tris boric acid, and 2ml of 0.5M EDTA) with 0.2μl ethidium as a fluorescent tag. The expression products were visualized as bands (amplicons) using UV-transilluminator. Snapshots of amplicons (figure 1) were densitometrically analyzed using ImageJ software (1.49V). Relative expression of miRNAs and p53 gene were calculated following endogenous normalization using miR-16 for miRNAs and β-actin for the p53 gene).^9,10^

**Figure 1:**
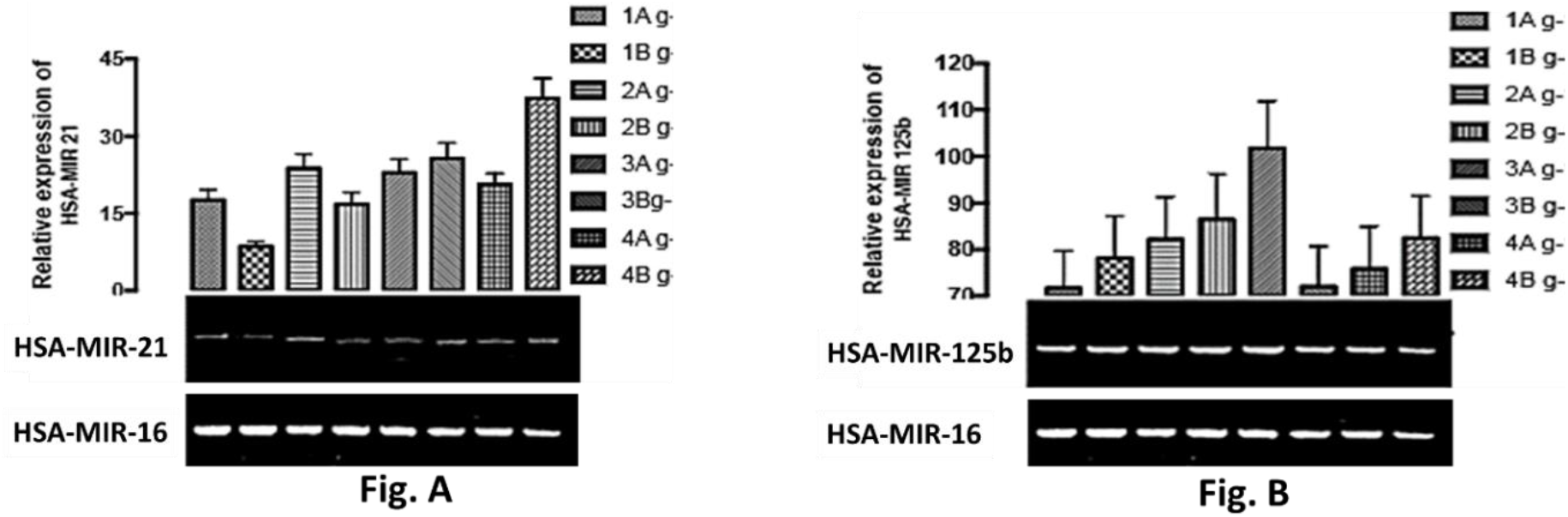
Gel electrophoretic image and expression of miR-21 (Fig. A) and miR-125b (Fig. B)

### Statistical analyses

The relative expression of miRNAs and p53 from two groups: Group 1 (HIV uninfected women) and Group 2 (Women living with HIV) were compared using the T-test and Pearson’s (bivariate) correlation using SPSS (version 22). Results were presented as mean ± standard error of mean (SEM). Statistical significance was set at p≤ 0.05.

## Results

The mean age of HIV-women (38.33 ± 10.04 years) was insignificantly lower compared with HIV+ women (41.36 ± 10.23 years) at p= 0.290. The CD4+ T-cell counts were insignificantly higher among HIV-women (1690 cells/μL) compared with HIV+ women (763.1 cells/μL) at p= 0.055.

### Levels of Oncogenes and tumour suppressors in serum

In serum samples (Figures 2C and 2D), a higher level of miR-21 was observed among HIV+ women compared with HIV-women (p< 0.05) while lower levels of miR-146a, miR-155, miR-182, and miR-200c were observed among HIV+ women compared with HIV-women at p< 0.05, p> 0.05, p> 0.05, and p> 0.05, respectively (Fig 2C). Also, a higher level of miR-145 was observed among HIV+ women compared with HIV-women (p> 0.05) while lower levels of miR-let-7b, miR-125b, miR-143, and p53 gene were observed among HIV+ women compared with HIV-women at p> 0.05, p= 0.05, p> 0.05, and p< 0.05, respectively (Fig. 2D). The low levels of miR-let-7b, miR-125b, miR-143, and p53 among HIV+ women could be due to immune exhaustion.

**Figure 2:**
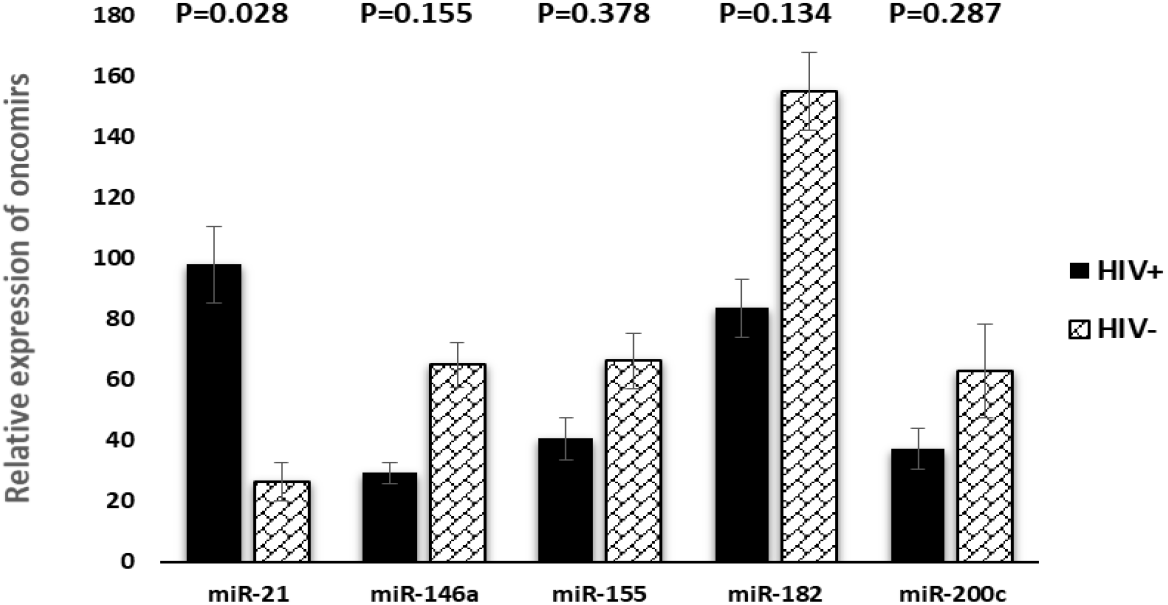
Relative expression of oncomirs in serum of HIV+ and HIV- women.

**Figure 3:**
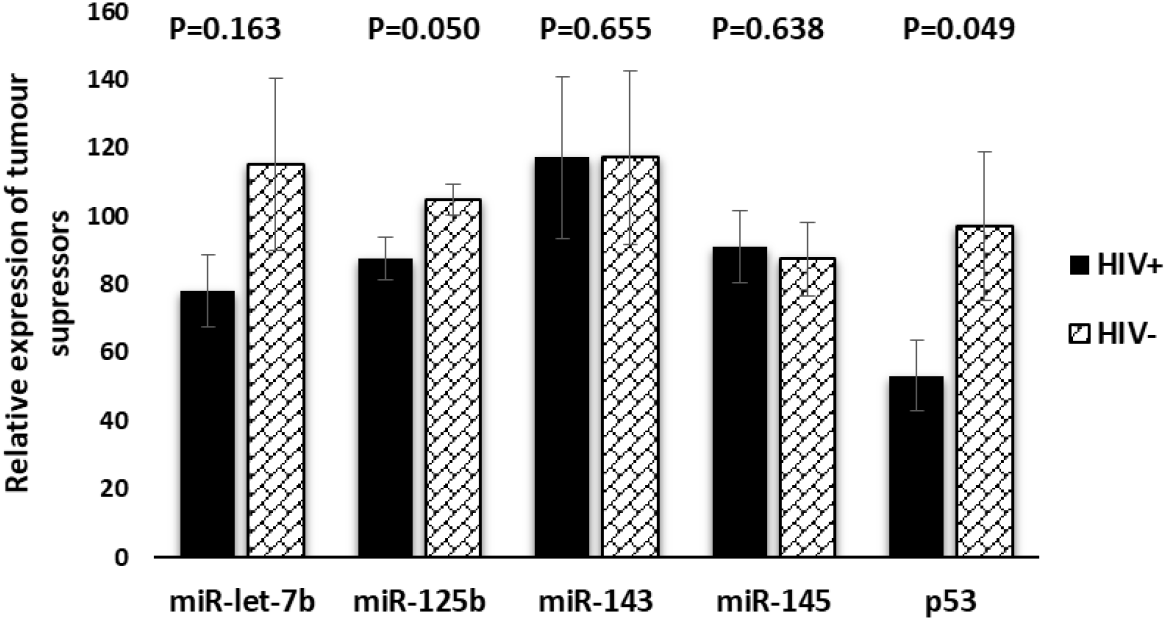
Relative expression of tumour suppressors in serum of HIV+ and HIV- women].

### Relationships between tumour suppressors and oncomirs

In the serum of HIV+ and HIV-women, significant direct relationships were between miR-21 and miR-155 (p= 0.002 and p= 0.014), miR-21 and 182 (p= 0.041 and p= 0.045, respectively) while a significant inverse relationship was observed between miR-21 and miR-let-7b (p= 0.013 and p= 0.049, respectively). Among HIV+ women, significant inverse relationships were observed between miR-200c and miR-let-7b (p= 0.039), and miR-let-7b and miR-145 (p= 0.006), while a direct relationship was observed between miR-155 and miR-145 (p= 0.049), miR-155 and miR-182 (p= 0.001), and miR-21 and miR-200c (p= 0.048). Among HIV-women, significant direct relationships were observed between miR-146a and miR-145 (p= 0.033), miR-146a and p53 (p= 0.030), miR-155 and miR-200c (p= 0.006), miR-155 and p53 (p= 0.012), and miR-200c and p53 gene (p= 0.003).

## Discussion

In literature, a higher peripheral blood level of miR-21 was observed among HIV+ individuals compared with their HIV-counterparts.^12^ This is consistent with the findings of this study and could be due to sustained stimulation, dysfunction, and apoptosis of CD4+ T cells among HIV+ women.^13^ It could also be due to a low level of circulating miR-let-7b considering the fact that there was an inverse relationship between miR-21 and miR-let-7b.^8^ Thus, miR-21 could be used as an early indicator of oncoviral activity and epithelial transformation among HIV+ women. Likewise, in this study, a significantly lower level of miR-125b was observed in the serum of HIV+ women compared with their HIV-counterparts. This could be due to immunosuppression following enhanced HIV replication,^14^ or CD4+ T cell activation.^15^ This suggests that the level of miR-125b could also be used to monitor immunosuppression-dependent epithelial transformation associated with HIV infection, irrespective of CD4+ T-cell counts and the nature of the sample.

An earlier study carried out among individuals living with HIV-1 reported a significantly higher plasma level of miR-155 among highly-active ART (HAART) naïve participants and non-responders compared with HAART responders and uninfected participants.^16^ In this study, there was an insignificant lower serum level of miR-155 among HIV+ women compared with HIV-women. The variation between the findings of Jin et al. and the findings of this study could be due to the nature of the sample assayed; plasma versus serum. MiR-155 is produced not only by mononuclear cells but also by CD4+ and CD8+ T lymphocytes found in plasma.^16^ The cells are abundant in the microenvironment of the cervical epithelium. Researchers have shown that infection with Epstein-Barr virus (EBV) infection results in cellular expression of miR-155.^17^. On the other hand, since the HIV+ women were serologically negative for EBV and HAART-experienced, the lower miR-155 among them suggests that they were majorly HAART responders. In this study, the low serum level of wild type p53 gene among HIV+ women is in consonance with the findings of Akwiwu et al. who reported a lower level of serum p53 protein and CD4+ T cell counts among HAART naïve HIV+ women followed by HAART-experienced HIV+ women compared with uninfected women.^18^ This suggests that HIV promotes p53 degradation or mutation, hence the low level in HIV+ women.

Parikh et al. reported a higher level of miR-145 in HIV+ individuals compared with HIV-counterparts. Despite the fact that the majority of the participants in their study were on anti-retroviral therapy (ART), the mean CD4+ T-cell counts of their participants were 484 cells/μL.^12^ In this study, although miR-145 was slightly higher among HIV+ women compared with HIV-women, no significant difference was observed between the two groups. It could be argued that the minimal differences in the expression of tumor suppressors between the groups could be due to the responsiveness of the HIV+ women to HAART. This is underscored by the fact that the CD4+ T-cell counts among the HIV+ women in this study are higher compared with the CD4+ T-cell count among the HIV+ participants in the study carried out by Parikh et al.^12^ In humans, miR-143 exerts anti-inflammatory effects and its low serum level is predictive of poor prognosis among critically ill patients.^19,20^ Since inflammation precedes and co-exists with cancer, the level of miR-143 among HIV+ women suggests that they are at low risk of developing cervical cancer.

Egana-Gorrono et al. and Nahand et al. reported higher levels of miR146a and miR-200c in mononuclear cells of viraemic HIV+ individuals, especially treatment naïve individuals, compared with uninfected individuals and elite controllers (ART naïve individuals with < 50 HIV-1 RNA copies/mL and CD4 T cell count of 200 to1000 cells/lL).^21,22^ Thus, the low levels of miR-146a, miR-182, and miR-200c in the serum of HIV+ women could be due to viral latency, reduced viral load, or low rate of T cell reactivation. Interestingly, viral latency forestalls HIV elimination in reservoir cells among HAART-experienced persons.^7,23^

## Limitations

The viral load in blood was not determined. This would have revealed the modulatory effects of HIV replication on the oncogenes and tumour suppressors.

## Conclusion

This study revealed a high circulating level of miR-21 and a low circulating level of miR-125b among HIV+ women in Southern Nigeria. The biomarkers could be used in identifying individuals at risk of developing cervical cancer, especially among immunocompromised patients. More so, developing a drug that can downregulate miR-21 and upregulate miR-125b may forestall malignant epithelial transformation among HIV+ women.

## Data Availability

All data produced in the present study are available upon reasonable request to the authors

## Acknowledgements

The authors owe a great deal of gratitude to the staff members of the Centre for Biocomputing and Drug Development, Adekunle Ajasin University, and HIV counseling and testing and family planning clinics, State Hospital Abeokuta, for their technical assistance.

## Data Availability Statement

Data will be made available on reasonable request.

## References

1. Sung H, Ferlay J, Siegel RL, Laversanne M, Soerjomataram I, Jemal A, Bray F. Global cancer statistics 2020: GLOBOCAN estimates of incidence and mortality worldwide for 36 cancers in 185 countries. CA: a Cancer J Clin. 2021 May;71(3):209–49.

2. Awofala AA, Ogundele OE. HIV epidemiology in Nigeria. Saudi J Biol Sci. 2018 May; 25 (4): 697–703.

3. Adewole IF, Benedet JL, Crain BT, Follen M. Evolving a strategic approach to cervical cancer control in Africa. Gynecol Oncol. 2005 Dec 1;99(3):S209–12.

4. Odutola M, Jedy-Agba EE, Dareng EO, Oga EA, Igbinoba F, Otu T, Ezeome E, et al. Burden of cancers attributable to infectious agents in Nigeria: 2012–2014. Front Oncol. 2016 Oct 24;6:216.

5. Denslow SA, Rositch AF, Firnhaber C, Ting J, Smith JS. Incidence and progression of cervical lesions in women with HIV: a systematic global review. Int J STD & AIDS. 2014 Mar;25(3):163–77.

6. Lutz G, Jurak I, Kim ET, Kim JY, Hackenberg M, Leader A, et al. Viral ubiquitin ligase stimulates selective host microRNA expression by targeting ZEB transcriptional repressors. Viruses. 2017 Aug 7;9(8):210.

7. Suzuki K, Matsubara H. Recent advances in p53 research and cancer treatment. Journal of Biomed Biotechnol. 2011 Jul;2011.

8. Okoye JO, Ngokere AA, Onyenekwe CC, Ogenyi SI, Omotuyi O. Circulating mir21 and mir125b in women living with human immunodeficiency virus: Utility of biomarkers for monitoring cervical carcinogenesis. Cancer Res. 2022 Jun 15;82(12_Supplement):1496-.

9. Okoye JO, Ngokere AA, Onyenekwe CC, Omotuyi O, Dada DI. Epstein-Barr virus, human papillomavirus and herpes simplex virus 2 co-presence severely dysregulates miRNA expression. Afr J Lab Med. 2021;10(1):1–0

10. Okoye JO, Ngokere AA, Onyenekwe CC, Erinle CA. Comparable expression of miR-let-7b, miR-21, miR-182, miR-145, and p53 in serum and cervical cells: Diagnostic implications for early detection of cervical lesions. Int J Health Sci. 2019 Jul;13(4):29.

11. Toni LS, Garcia AM, Jeffrey DA, Jiang X, Stauffer BL, Miyamoto SD, Sucharov CC. Optimization of phenol-chloroform RNA extraction. MethodsX. 2018 Jan 1;5:599–608.

12. Parikh VN, Park J, Nikolic I, Channick R, Paul BY, De Marco T, et al. Coordinated modulation of circulating miR-21 in HIV, HIV-associated pulmonary arterial hypertension, and HIV/HCV coinfection. J acquir Immune Defic Syndr (1999). 2015 Nov 11;70(3):236.

13. Nguyen LN, Nguyen LN, Zhao J, Schank M, Dang X, Cao D, et al. Long non-coding RNA GAS5 regulates T cell functions via miR21-mediated signaling in people living with HIV. Front Immunol. 2021 Mar 12;12:601298.

14. Jin C, Cheng L, Höxtermann S, Xie T, Lu X, Wu H, et al. Micro RNA-155 is a biomarker of T-cell activation and immune dysfunction in HIV-1-infected patients. HIV Med. 2017 May;18(5):354–62.

15. Mantri CK, Pandhare Dash J, Mantri JV, Dash CC. Cocaine enhances HIV-1 replication in CD4+ T cells by down-regulating MiR-125b. PloS one. 2012 Dec 12;7(12):e51387.

16. Ruelas DS, Chan JK, Oh E, Heidersbach AJ, Hebbeler AM, Chavez L, et al. MicroRNA-155 reinforces HIV latency. J Biol Chem. 2015 May 29;290(22):13736–48.

17. Egaña-Gorroño L, Escribà T, Boulanger N, Guardo AC, León A, Bargalló ME, et al. HIV Controllers Consortium of the AIDS Spanish Network. Differential microRNA expression profile between stimulated PBMCs from HIV-1 infected elite controllers and viremic progressors. PLoS One. 2014 Sep 16;9(9):e106360.

18. Sadri Nahand J, Bokharaei-Salim F, Karimzadeh M, Moghoofei M, Karampoor S, Mirzaei HR, et al. MicroRNAs and exosomes: key players in HIV pathogenesis. HIV Med. 2020 Apr;21(4):246–78.

19. Linnstaedt SD, Gottwein E, Skalsky RL, Luftig MA, Cullen BR. Virally induced cellular microRNA miR-155 plays a key role in B-cell immortalization by Epstein-Barr virus. J Virol. 2010 Nov 15;84(22):11670–8.

20. Huang J, Wang F, Argyris E, Chen K, Liang Z, Tian H, et al. Cellular microRNAs contribute to HIV-1 latency in resting primary CD4+ T lymphocytes. Nature Med. 2007 Oct;13(10):1241–7.

21. Li SY, Zhang ZN, Jiang YJ, Fu YJ, Shang H. Transcriptional insights into the CD8+ T cell response in mono-HIV and HCV infection. J Trans Med. 2020 Dec;18(1):1–1

22. Roderburg C, Koch A, Benz F, Vucur M, Spehlmann M, Loosen SH, et al. Serum levels of miR-143 predict survival in critically ill patients. Disease markers. 2019 Oct 23;2019.

23. Akwiwu E, Okafor A, Akpan P, Akpotuzor J, Asemota E, Okoroiwu H, Anyanwu S. Serum P53 Protein Level and Some Haematologic Parameters among Women of Reproductive Age Living with HIV Infection. Nig J Physiol Sci. 2021 Jun 30;36(1):85–9.

